# SARS-CoV-2 natural antibody response persists up to 12 months in a nationwide study from the Faroe Islands

**DOI:** 10.1101/2021.04.19.21255720

**Authors:** Maria Skaalum Petersen, Cecilie Bo Hansen, Marnar Fríðheim Kristiansen, Jógvan Páll Fjallsbak, Sólrun Larsen, Jóhanna Ljósá Hansen, Ida Jarlhelt, Laura Pérez-Alós, Bjarni á Steig, Debes Hammershaimb Christiansen, Lars Fodgaard Møller, Marin Strøm, Guðrið Andorsdóttir, Shahin Gaini, Pál Weihe, Peter Garred

## Abstract

Only a few studies have assessed the long-term duration of the humoral immune response against severe acute respiratory syndrome coronavirus 2 (SARS-CoV-2).

In this nationwide longitudinal study from the Faroe Islands with close to full participation of all individuals on the Islands with PCR confirmed COVID-19 during the two waves of infections in the spring and autumn 2020 (n=172 & n=233), samples were drawn at three longitudinal time points (3, 7 and 12 months and 1, 3 and 7 months after disease onset, respectively).

Serum was analyzed with a direct quantitative IgG antibody binding ELISA to detect anti–SARS-CoV-2 spike RBD antibodies and a commercially available qualitative sandwich RBD ELISA kit measuring total antibody binding.

The seropositive rate in the convalescent individuals was above 95 % at all sampling time points for both assays. There was an overall decline in IgG titers over time in both waves (p < 0.001). Pairwise comparison showed that IgG declined significantly from the first sample until approximately 7 months in both waves (p < 0.001). After that, the antibody level still declined significantly (p < 0.001), but decelerated with an altered slope remaining fairly stable from 7 months to 12 months after infection. Interestingly, the IgG titers followed a U-shaped curve with higher antibody levels among the oldest (67+) and the youngest (0– 17) age groups compared to intermediate groups (p < 0.001).

Our results indicate that COVID-19 convalescent individuals are likely to be protected from reinfection up to 12 months after symptom onset and maybe even longer. We believe our results can add to the understanding of natural immunity and the expected durability of SARS-CoV-2 vaccine immune responses.

## Research letter

Only a few studies have assessed the long-term duration of the humoral immune response against severe acute respiratory syndrome coronavirus 2 (SARS-CoV-2). Recently, an American study found that IgG titers were durable, with modest declines in titers 6 to 8 months after symptom onset^1^ while a Canadian study showed a reduction in circulating antibodies over time but increasing and persisting levels of receptor-binding domain (RBD)-specific B cells up to 8 months after symptom onset^2^. Here, we report results of natural IgG response to SARS-CoV-2 infection up to 12 months from symptom onset in a longitudinal nationwide study from the Faroe Islands^3^ with participation of more than 80% of all individuals with polymerase chain reaction (PCR) confirmed COVID-19 during two waves of infections (March to April and August to December 2020). Samples were drawn at three longitudinal time points for each wave: wave 1 at median 89, 210 and 363 days (range 29–380 days), and wave 2 at median 27, 125 and 210 days (range 27– 231 days) after symptom onset, respectively (wave 1: n = 172, 54% women, median age 42, range 1–93 years; wave 2: n = 233, 53% women, median age 35, range 0–83 years). There was no observed overlap among the two waves, indicating that wave 1 participants were not reinfected in wave 2. The disease course ranged from asymptomatic to critically ill, with only 6 and 8 individuals hospitalized, respectively (table 1 for study characteristics).

**Table 1.** Characteristics of the COVID-19 patients in the Faroe Islands from March 2020 to December 2020, stratified in two waves (n = 172 and n = 233)

|  | Wave 1 (n = 172) |  | Wave 2 (n = 233) |  |
| --- | --- | --- | --- | --- |
| **Sex, n (%)** |  |  |  |  |
| Female | 92 | 53.5 | 123 | 52.8 |
| **Age** |  |  |  |  |
| Age (years), median (range) | 42.3 | 1.2–92.9 | 34.7 | 0.27–82.6 |
| Age distribution, n (%) |  |  |  |  |
| 0-17 | 17 | 9.8 | 27 | 11.6 |
| 18-34 | 54 | 31.2 | 90 | 38.6 |
| 35-49 | 39 | 22.5 | 56 | 24.0 |
| 50-66 | 45 | 26 | 41 | 17.6 |
| 67+ | 18 | 10.4 | 19 | 8.2 |
| **Symptoms during the acute phase<sup>b</sup>, n (%)** |  |  |  |  |
| Asymptomatic<sup>a</sup> | 33 | 19.2 | 32 | 15.9 |
| Cough * | 78 | 45.3 | 116 | 57.4 |
| Fever* | 110 | 64.0 | 107 | 53.2 |
| Sore throat* | 53 | 30.8 | 77 | 38.3 |
| Headache* | 84 | 48.8 | 112 | 60.7 |
| Dyspnea* | 19 | 11.0 | 32 | 15.9 |
| **Days from symptoms onset to blood sampling, median (range)** |  |  |  |  |
| First blood sample | 89 | 29–163 | 27 | 10–79 |
| Second blood sample | 210 | 171–294 | 125 | 45–191 |
| Third blood sample | 363 | 302–380 | 210 | 94–231 |
| **Number of samples delivered, n** |  |  |  |  |
| First sample | 168 |  | 221 |  |
| Two sample | 139 |  | 221 |  |
| Three samples | 158 |  | 153 |  |
| **Disease groups, n (%)<sup>c</sup>** |  |  |  |  |
| Hospitalized | 8 | 4.7 | 6 | 3.0 |
| Asymptomatic<sup>a</sup> | 33 | 18.6 | 31 | 15.3 |
| Symptomatic | 132 | 76.7 | 165 | 81.7 |

<sup>a</sup>Asymptomatic for the five symptoms which were asked about directly, not necessarily other symptoms; <sup>b</sup>n = 32 missing in wave 2;
<sup>c</sup>n = 31 missing in wave 2.

Serum was analyzed with a direct quantitative IgG antibody binding ELISA to detect anti–SARS-CoV-2 spike RBD antibodies^4^ and a commercially available qualitative sandwich RBD ELISA kit measuring total antibody binding (Beijing Wantai Biological Pharmacy Enterprise Co., Ltd., China).

The seropositive rate in the convalescent individuals was above 95% at all sampling time points for both assays and remained stable over time, i.e. almost all convalescent individuals developed antibodies. There was an overall decline in IgG titers over time in both waves (Friedman test: χ^2^(2) = 133.273, *p* < 0.001 and χ^2^(2) = 112.407, *p* < 0.001) (figure 1a). Wilcox Signed-Ranks pairwise comparison showed that IgG declined significantly from the first sample until approximately 7 months in both waves (*p* < 0.001; figure 1a). After that, the antibody level still declined significantly, but decelerated with an altered slope remaining fairly stable from 7 months to 12 months after infection (*p* < 0.001). Antibody levels did not differ significantly between women and men over time (*p* = 0.09 – *p* = 0.7). Interestingly, the IgG titers followed a U-shaped curve with higher antibody levels among the oldest (67+) and the youngest (0–17) age groups compared to intermediate groups (figure 1b and 1c) (Kruskal-Wallis test: χ^2^(4) = 15.132, *p* = 0.004 and χ^2^(4) = 52.582, *p* < 0.001).

**Figure 1.**
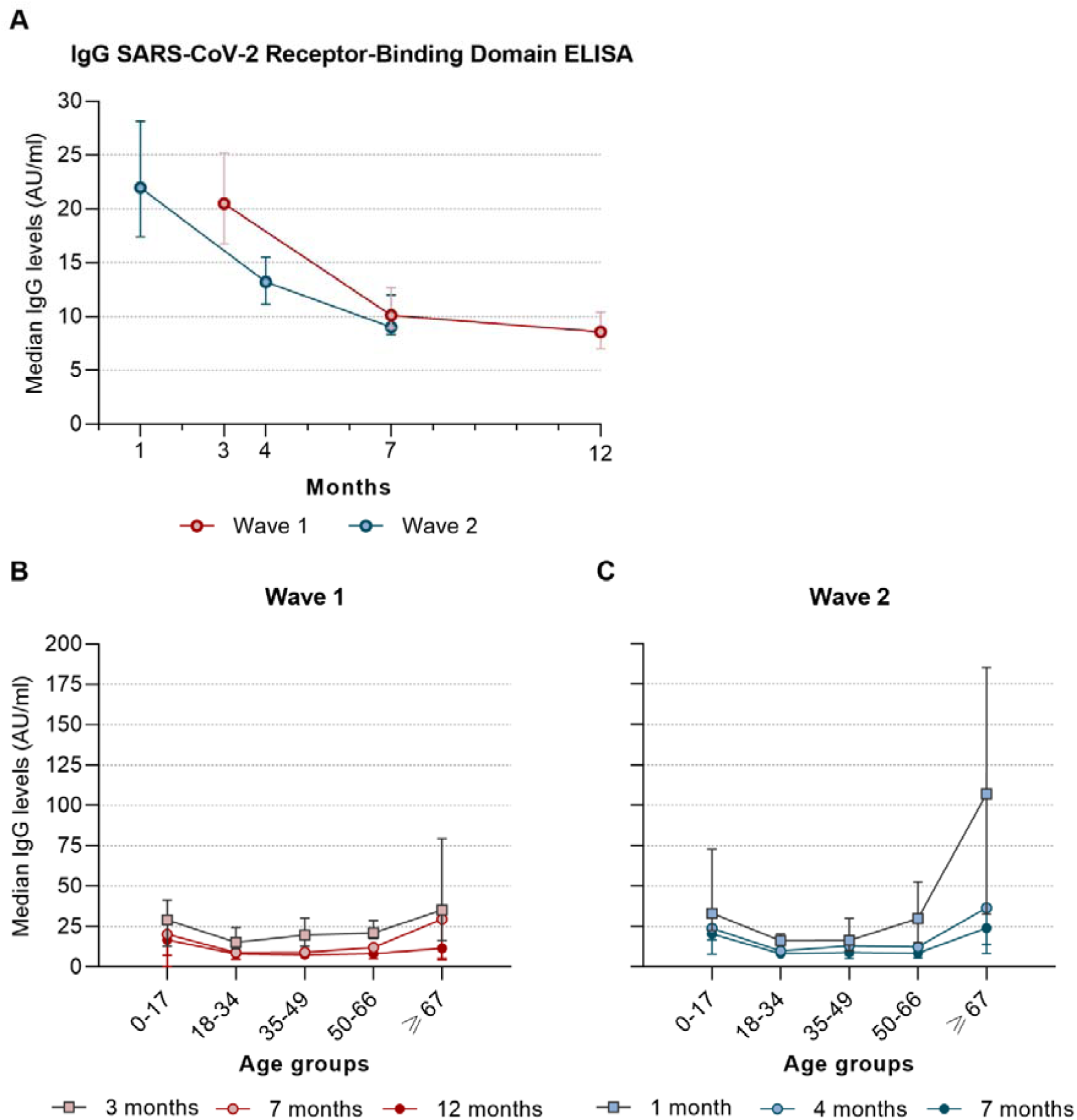
Longitudinal analyses of serum samples from COVID-19 patients in the Faroe Islands from March 2020 to December 2020, stratified in two waves. **a)** Median IgG antibody titers plotted against median time from symptom onset to sampling. Data is expressed as median with 95% confidence intervals. Median antibody titers were 20.5 AU/ml (range:0–584.3 AU/ml), 10.1 AU/ml (0–200 AU/ml) and 8.6 AU/ml (0–104.7 AU/ml) respectively from 3 to up to 12 months in wave 1 (red line), and 22.0 AU/ml (0–842.5 AU/ml), 13.2 AU/ml (0– 719.4 AU/ml) and 9.0 AU/ml (0–388.4 AU/ml) from 1 to 7 months (blue line). Friedman test showed an overall decline in IgG titers over time in both waves (χ^2^(2) = 133.273, *p* < 0.001 and χ^2^(2) = 112.407, *p* < 0.001). Wilcox Signed-Ranks test pairwise comparison showed a significant decline between all time points (*p* < 0.001). **b)** Median IgG titers in wave 1 plotted against age group. Kruskal-Wallis test showed an overall significant difference in antibody levels in age groups (χ2(4) = 15.132, *p* = 0.001). Pairwise comparisons (Mann-Whitney U test) for each time point indicated that the oldest age group had higher median IgG titers compared with the age groups 18-34 years, albeit only significantly in the first and second sample (*p* = 0.03 and *p* = 0.05) while the youngest age group had significantly higher IgG levels compared with the age group 35-49 in third sample (*p* = 0.03). **c)** Median IgG titers in wave 2 plotted against on age group. Kruskal-Wallis test showed an overall significant difference in antibody levels in age groups (χ2(4) = 58.582, *p* < 0.001). Pairwise comparisons (Mann-Whitney U test) for each time point indicated that the oldest age group had significantly higher median IgG titers compared with all age groups (*p* ≤ 0.02) in first sample while in the second and third sample, IgG levels were higher compared with all age groups (*p* ≤ 0.009) except the youngest age group (*p* = 0.1 and *p* = 0.6). The youngest age group had significantly higher IgG levels compared with age groups 18-49 in the first and second sample (*p* ≤ 0.02 and *p* ≤ 0.01) and age group 18-66 in the third sample (*p* ≤ 0.02).

One limitation of the study is that we do not measure neutralizing antibodies. However, the neutralizing antibody and ELISA results generally correlate with each other^5^ and RBD is the target of most neutralizing antibodies against SARS-CoV-2^6^.

Albeit the protective role of antibodies is currently unknown, our results indicate that COVID-19 convalescent individuals are likely to be protected from reinfection up to 12 months after symptom onset and maybe even longer. Our results represent SARS-CoV-2 antibody immunity in nationwide cohorts in a setting with few undetected cases^7^ and we believe our results can add to the understanding of natural immunity and the expected durability of SARS-CoV-2 vaccine immune responses. Moreover, they can help in public health policies and ongoing strategies for vaccine delivery.

## Data Availability

The data is not public available but may be made available from the corresponding authors
upon reasonable request.

